# geneXplore: An Interactive Browser for X Chromosome-Wide Association Study Results

**DOI:** 10.64898/2026.07.14.26357489

**Authors:** Noah Cook, Jordan Boulais-Richard, Youjie Zeng, Chenyu Yang, John Budde, Daniel Taliun, Sarah A. Gagliano Taliun, Carlos Cruchaga, Michael E. Belloy

## Abstract

**Summary:** The X chromosome comprises approximately 5% of the human genome and encodes over 800 protein-coding genes, many of which exhibit sex-differentiated expression patterns due to escape from X chromosome inactivation (XCI) mechanisms. Despite its relevance to sex differences in complex traits, the X chromosome is routinely excluded from genome-wide association studies due to analytical challenges, and when analyzed, the impact of escape from XCI or sex is limitedly explored. No dedicated, publicly accessible browser for X chromosome-wide association study (XWAS) summary statistics currently exists, creating a barrier to systematic investigation of X-linked contributions to human traits. Here, we present geneXplore, an interactive web browser based on the PheWeb2 implementation, tailored for XWAS summary statistics across 1,944 phenotypes while distinguishing random XCI (rXCI), escape from XCI (eXCI), and sex-stratified analyses. Users can explore results via interactive plots (Manhattan and Miami, PheWAS and LocusZoom), searchable tables and access to cross-database lookup, with full summary statistics available for download.

**Availability and Implementation:** geneXplore is freely available at https://genexplore.wustl.edu/ with no registration required and will be maintained for a minimum of two years following publication. Source code is available at https://github.com/Belloy-Lab/geneXplore_XWAS_Browser under an MIT license.

## Introduction

Research on X chromosome genetics in humans lingers behinds the autosomes due to increased data processing and analytical complexities. Yet, this chromosome comprises approximately 5% of the total genome and encodes over 800 protein-coding genes, many of which are expressed in a sex-differentiated matter. While one X chromosome in females randomly undergoes X chromosome inactivation (XCI) to balance gene expression with males, many X-linked genes still show sex-differences expression due to (variable) escape from X chromosome inactivation (eXCI) in females that happens for over 30% of genes (Tukiainen et al. 2017, Sidorenko et al. 2019). This biological complexity, among other reasons, has led to the X chromosome being routinely excluded from standard genome-wide association studies (GWAS), which systematically test genetic variants across the genome for statistical association with a trait or disease (Gorlov and Amos 2023, Sun *et al*. 2023). When X chromosome analyses are performed, they commonly collapse biological heterogeneity by ignoring XCI status or include sex as a covariate in models that analyze males and females together (Sidorenko *et al*. 2019, Chen *et al*. 2021).

This analytical gap has important consequences on our understanding of the genetics of complex human conditions given that sex differences are widely observed in the prevalence, severity, age of onset, and pathobiology, for a wide range of complex traits, including Alzheimer’s disease (Belloy *et al*. 2024, Cook *et al*. 2025), autoimmune disorders (Forsyth *et al*. 2024), cardiovascular disease (Tcheandjieu *et al*. 2022), where X-linked genetic variants are expected to contribute meaningfully to these sex differences. Similarly, multiple studies have shown sex differences in the genetic regulation of gene expression that could in turn shape sex differences in disease etiology. Crucially, a dedicated, publicly accessible resource for X chromosome-wide association study (XWAS) data does not currently exist, creating a barrier to systematic investigation of X-linked contributions to human disease.

To address this gap, we developed geneXplore, a web-based XWAS browser, based on the PheWeb2 implementation (Bellavance *et al*. 2026), to provide open, interactive access to a curated collection of XWAS summary statistics across traits, ancestries, and XCI-informed analytical models. By lowering the barrier to exploring X chromosome data, geneXplore aims to accelerate discovery in this underserved region of the genome.

## Methods

### Data Content

geneXplore currently hosts XWAS summary statistics for 1,944 unique phenotypes. These mostly include the non-pseudoautosomal regions (non-PAR) but also the pseudoautosomal (PAR) regions when available, and are drawn from datasets across three trait categories:

1. **Dichotomous traits** (16 phenotypes) encompass neurodegenerative and cardiovascular diseases, including Alzheimer’s disease (Le Borgne et al. 2025, Cook et al. 2026), Parkinson’s disease (Le Guen et al. 2021), Lewy body dementia (Bayram *et al*. 2024), multiple system atrophy (Ray *et al*. 2026), coronary artery disease (Tcheandjieu *et al*. 2022), mosaic X chromosome loss in females (Liu *et al*. 2024), and dementia risk factor traits derived from UK Biobank summary statistics (Livingston *et al*. 2024).
2. **Continuous traits** (888 phenotypes) span neuroimaging, hormonal, lipid, and cognitive domains, including structural magnetic resonance imaging (sMRI) and diffusion tensor imaging (DTI) brain imaging traits (Jiang *et al*. 2025), blood lipid traits across five ancestries (Graham *et al*. 2021), blood sex hormone levels (Ruth *et al*. 2020), age at menarche (Kentistou *et al*. 2024), age at menopause (Ruth *et al*. 2021), cognitive resilience (Eissman *et al*. 2022), baseline memory and memory decline (Eissman *et al*. 2024), and mosaic X chromosome loss (Liu *et al*. 2024).
3. **Brain single-cell trans-expression quantitative trait loci** (sc-trans-eQTLs; 7 cell types; 1040 phenotypes) represent X chromosome variants associated with the expression of X chromosome genes across seven brain cell types, restricted to genes for which significant QTLs were observed in at least one cell type in the original study (Zeng *et al*. 2026, *in preparation*).

All XWAS summary statistics were mapped to the GRCh38 reference genome via liftover and processed through a standardized preprocessing pipeline.

### X Chromosome Genotype Modeling

A key feature of geneXplore is its explicit representation of X chromosome genotype modeling strategies. Specifically, geneXplore hosts association results for up to four analytical strata per phenotype and ancestry group. The four strata are: (1) a random XCI (rXCI) model, in which females allele dosages are coded as 0/1/2 and males as 0/2, assuming random inactivation of one X chromosome copy in females; (2) an escape from XCI (eXCI) model, in which females are coded as 0/1/2 and males as 0/1, assuming both X chromosome copies are active in females; (3) a female-stratified analysis restricted to female participants; and (4) a male-stratified analysis restricted to male participants.

This enables users to assess potential sex differences or biases toward eXCI or rXCI for genetic associations within and across traits. When available, eXCI summary statistics were derived from prior studies in which individual-level genotype data were directly encoded to assume eXCI. In other instances, eXCI summary statistics were derived via random-effects meta-analysis of male (coded as 0/1) and female (coded as 0/1/2) strata (Cook *et al*. 2026). Example code for both modeling approaches are available in the geneXplore GitHub repository.

### Browser Interface and Visualization

geneXplore is built on the PheWeb2 framework and PheWeb2-API (Bellavance *et al*. 2026), a Python-based backend that preprocesses and indexes summary statistics and serves them via a REST interface, with a Vue.js application frontend. PheWeb2 natively supports stratified GWAS result files, multi-stratum dropdown selection, and Manhattan/Miami, LocusZoom, and PheWas plot visualizations. geneXplore extends this framework with several X chromosome-specific modifications: decreased chromosomal binning to a 30 kb bin length to accommodate a single chromosome view, further modifications to improve visualization of X chromosome specific findings, and an X chromosome-wide significance threshold of P<1×10^−5^. This lower significance threshold is relevant since XWAS have lower burden of multiple comparisons and even more importantly because they have reduced power relative to autosomal GWAS (Gorlov and Amos 2023, Sun *et al*. 2023). This is because in women, genetic loci that undergo rXCI, have reduced impact on phenotypic variance due to uncertainty of the active genotype, reducing power by 75%, while in men the single X chromosome copy, i.e. hemizygosity, reduces power by 50% (Sidorenko *et al*. 2019). The use of a more relaxed significance threshold is therefore conductive to accelerate new research and insights into the X chromosome.

## Results

geneXplore hosts association summary statistics across 1,944 unique phenotypes, spanning 6,917 stratum-by-phenotype combinations across up to four XCI-informed analytical models and up to six ancestry groups (**Figure.1A**). For European-ancestry analyses, the number of X chromosome variants tested per phenotype averages approximately 270,000, though this varies by source study and phenotype category, with some large-scale consortia datasets contributing substantially denser variant coverage. These variants span the full minor allele frequency (MAF) spectrum and are skewed toward rare and low-frequency variants, with 45% having MAF<1%, 16% having 1%≤MAF<5%, and 39% having MAF≥5%. In total, 981 phenotypes had at least one X chromosome-wide significant locus in at least one stratification (P<1×10^−5^).

**Figure 1.**
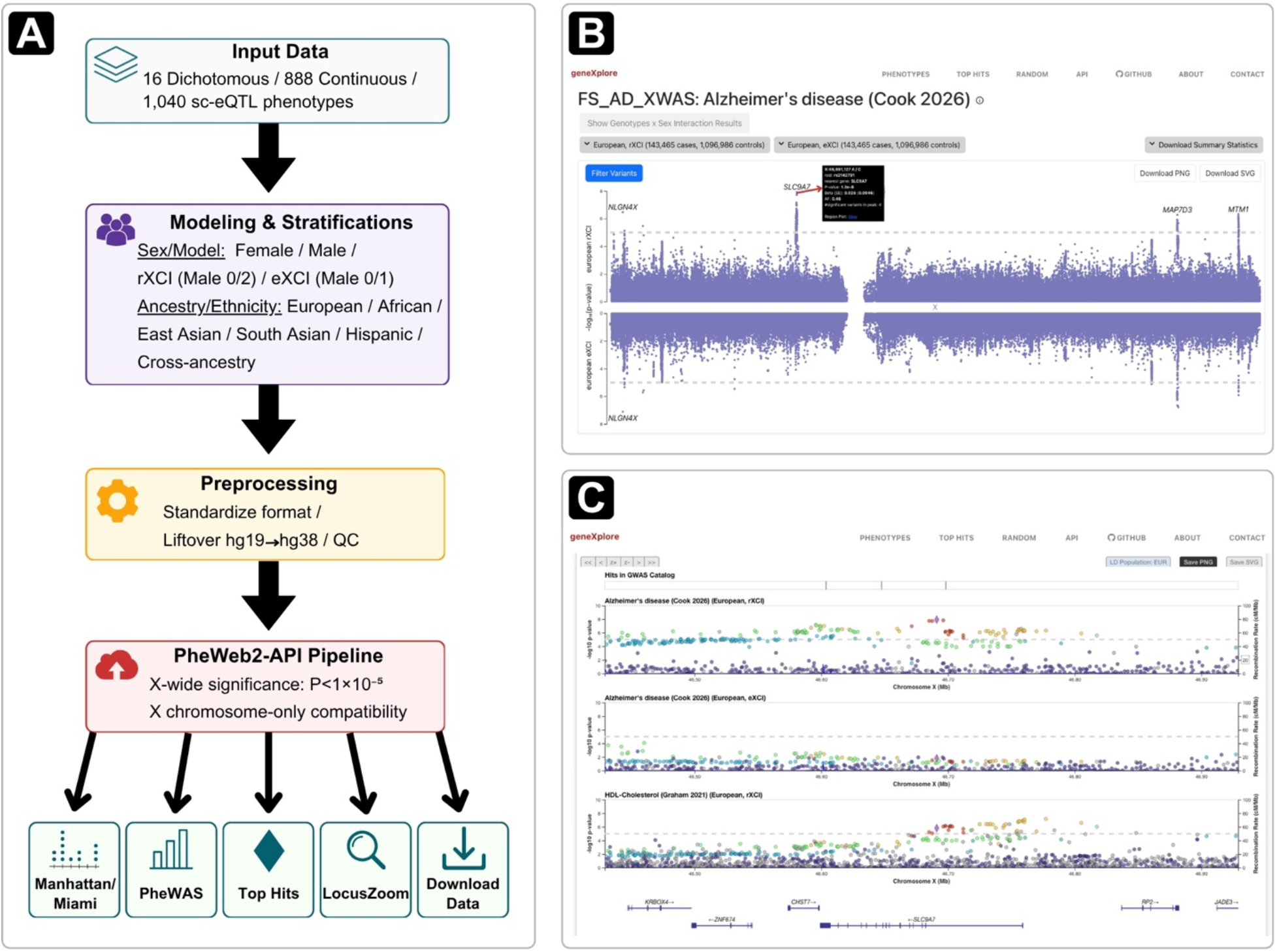
Overview of the geneXplore pipeline and browser functionality. **A)** Schematic of the geneXplore data processing pipeline and framework for X chromosome-tailored visualization using the PheWeb2-API pipeline. **B)** Miami plot view for Alzheimer’s disease, comparing European rXCI (top) and eXCI (bottom) models across the X chromosome. An interactive tooltip displays variant-level statistics for rs2142791 near *SLC9A7*, with direct links to external databases. **C)** Stacked regional association viewer (LocusZoom) at the *SLC9A7* locus, overlaying Alzheimer’s disease rXCI [top] and eXCI [middle] with HDL-Cholesterol rXCI [bottom]; linkage disequilibrium data is calculated from the European 1000 Genomes reference panel.

To demonstrate geneXplore’s functionality, we present a case example centered on an X-linked association near *SLC9A7* in Alzheimer’s disease (Cook *et al*. 2026). Navigating to the Alzheimer’s disease phenotype, the Miami plot view enables simultaneous comparison of two user-selected stratifications (**Figure.1B**). At the *SLC9A7* locus, an association signal is present under the rXCI model (P=1.2x10^-8^) but remains sub-threshold in the eXCI stratum (P=1.5x10^-2^), illustrating how the browser facilitates direct comparison across XCI modeling assumptions. Additional data exploration options allow to overlay LocusZoom plots (i.e. local visualizations of a genomic region of interest) across multiple phenotypes at a locus of interest simultaneously (**Figure.1C**). Comparing the Alzheimer’s disease lead variant (rs2142791) across Alzheimer’s disease rXCI (Cook *et al*. 2026), Alzheimer’s disease eXCI (Cook *et al*. 2026), and HDL-Cholesterol rXCI associations (Graham *et al*. 2021), reveals this genetic association likely follows an rXCI rather than an eXCI model and displays pleiotropy (i.e. influencing multiple distinct traits), across Alzheimer’s disease and HDL-cholesterol. Each locus is also annotated with variants previously reported as genome-wide significant in GWAS Catalog, enabling comparison against a broader array of phenotypes beyond those hosted in geneXplore. For example, at the *SLC9A7* locus, three previously reported genome-wide significant associations are displayed from a study not included in geneXplore: lymphocyte percentage of white cells, neutrophil count, and hemoglobin (Vuckovic *et al*. 2020), suggesting that this locus may have broader pleiotropic effects extending beyond Alzheimer’s disease and HDL-cholesterol. Finally, a complementary PheWAS view displays association statistics for any queried variant across all phenotypes and categories simultaneously, enabling screening of associations across all available stratifications.

## Discussion

GeneXplore is specifically designed around the biological and statistical complexities of the X chromosome, incorporating XCI-aware modeling, sex stratification, and X chromosome-adapted significance thresholding and visualization. While the current resource can be expanded with additional XWAS datasets that will become available, the tools and framework are directly shared to enable other researchers in building equivalent resources to share their XWAS results. One limitation, which applies to all web-browsers using publicly available datasets, is that our resource depends on the quality of the underlying summary statistics and researchers should therefore remain mindful of potential biases. Relatedly, coverage of the pseudoautosomal regions (PAR1 and PAR2) is not uniform across phenotypes, as it depends on whether these variants were retained in each source dataset. Most notably, an important consideration for the X chromosome is the inherent differential power across sexes that additionally depends on whether a locus displays eXCI for a given phenotype (Sidorenko *et al*. 2019). As such, users need to be attentive to the modelling assumptions and carefully evaluate not just association significance but also effect sizes across sexes in light of the reported genotype coding scheme.

In summary, geneXplore fills a critical gap by providing a dedicated, publicly accessible browser to visualize, compare and share XWAS summary statistics. Crucially, this resource will support researchers to investigate the role of the X chromosome in human health and disease, as well as its role in shaping sex differences.

## Data Availability

Summary statistics for all phenotypes hosted in geneXplore are available for download directly through the browser; details on data sources and availability by phenotype are provided on the About page at https://genexplore.wustl.edu/about.

## Author contributions

N.C.: Data curation, Formal analysis, Software, Visualization, Writing - original draft. J.B.R.: Software, Visualization. Y.Z.: Data curation. C.Y.: Visualization. J.B.: Software, Infrastructure. D.T. and S.A.G.T.: Methodology, Software, Writing – review & editing. C.C.: Infrastructure. M.E.B.: Funding acquisition, Supervision, Writing - review & editing.

## Code Availability

Codes for geneXplore are available at https://github.com/Belloy-Lab/geneXplore_XWAS_Browser. PheWeb2 and PheWeb2-API codes are respectively available at https://github.com/GaglianoTaliun-Lab/PheWeb2, and https://github.com/GaglianoTaliun-Lab/PheWeb2-API.

## Acknowledgements

The authors would like to thank Dan Western and Ellen Liu at Washington University Neurogenomics and Informatics Center for providing valuable feedback on the usability of the platform; the Washington University Medicine Marketing and Communications team for their contributions to the branding and public presentation of geneXplore; and Washington University Medicine Information Technology for hosting and infrastructure support.

SAGT and DT acknowledge support from the Canadian Institutes of Health Research (CIHR) (AD6-192920, PJT-197954 and AD7-200181). SAGT acknowledges salary support from a Fonds de Recherche du Québec – Santé (FRQS; https://frq.gouv.qc.ca) Junior 2 Award (https://doi.org/10.69777/347366). SAGT and DT acknowledge the support of the Natural Sciences and Engineering Research Council of Canada (NSERC) Discovery Grant Program: RGPIN-2026-04385 (SAGT) and RGPIN-2025-06572 (DT).

